# Automated chest radiograph diagnosis: A Twofer for Tuberculosis and Covid-19

**DOI:** 10.1101/2020.10.13.20178483

**Authors:** Mitushi Verma, Deepak Patkar, Madhura Ingalharikar, Amit Kharat, Pranav Ajmera, Viraj Kulkarni, Aniruddha Pant, Vaishnavi Thakker, Gunjan Naik

## Abstract

Coronavirus disease (Covid 19) and Tuberculosis (TB) are two challenges the world is facing. TB is a pandemic which has challenged mankind for ages and Covid 19 is a recent onset fast spreading pandemic. We study these two conditions with focus on Artificial Intelligence (AI) based imaging, the role of digital chest x-ray and utility of end to end platform to improve turnaround times. Using artificial intelligence assisted technology for triage and creation of structured radiology reports using an end to end platform can ensure quick diagnosis. Changing dynamics of TB screening in the times of Covid 19 pandemic have resulted in bottlenecks for TB diagnosis. The paper tries to outline two types of use cases, one is COVID-19 screening in a hospital-based scenario and the other is TB screening project in mobile van setting and discusses the learning of these models which have both used AI for prescreening and generating structured radiology reports.

## Introduction

Coronavirus disease 2019 (COVID-19) is a highly infectious disease caused by severe acute respiratory syndrome coronavirus [1]. The outbreak of Covid-19 originated in Wuhan, China in December 2019 and has been an ongoing pandemic since then across the world [2]. The disease quickly spread beyond China affecting the majority of the world with almost 37.8 million confirmed cases and **1**.**08 million** deaths as of October 13th, 2020, 13:03 [3]. The pandemic has brought the world to a standstill as most of the countries have been following a complete lockdown to curb the infection, where people isolate themselves at home and socialization of any type is discouraged.

Effective and speedy screening of Covid-19 infected patients is extremely crucial not only for isolating the patients to mitigate the spread of the virus but also to provide timely treatment with efficacy. To this end, contact tracing and containment have been undertaken with full force to identify positive cases and to curb community transmission. Despite all these measures, as the virus is novel, extensive studies have not yet been performed and the world still is looking for evidence-based medicine and sharing new guidelines on diagnostic testing [4]. Currently, clinical examination, complete blood count, digital chest x-ray, computed tomography (CT), real time reverse transcription–polymerase chain reaction (RT-PCR) and rapid antigen testing are leading to the diagnosis, with RT-PCR being the most popular test adopted globally. RT-PCR facilitates reliable diagnosis of Covid-19, nonetheless, is expensive, complicated as it involves a laborious process that not only requires PCR kits but also availability of expertise and sophisticated machinery in abundant locations across each country. To mitigate this, rapid antigen testing which provides the result within an hour, has been recently encouraged. However, this testing has a very high false negative rate and a follow up RT-PCR is required in majority cases. With an overwhelmingly increasing number of cases each day, that is already overburdening the health-care systems across the world, RT-PCR type time-consuming tests may not suffice and ancillary diagnostic solutions are imperative.

Chest imaging has been illustrated to be crucial in diagnosis and management of Covid-19, as the virus attacks the lungs causing pneumonia like etiology. To this end, the radiology guidelines suggest chest x-ray acquisition for infected patients with follow up scans for older, comorbid patients as well as for patients with worsening respiratory status and moderate-severe clinical symptoms. For majority patients the radiographs may illustrate airspace opacities either consolidations or ground glass opacities (GGO) even at an early stage [4]. The abnormalities are often bilateral, peripheral and prominent in the lower zone [5]. Moreover, the radiographic abnormalities may not necessarily correlate with the clinical symptoms. A study on 112 patients found that 54% patients demonstrated pneumonia-like changes on the radiograph however were completely asymptomatic [6]. Furthermore, the only risk factors into worsening of the disease that have been currently highlighted are older age, obesity and other comorbidities where the radiographs in severe cases might demonstrate acute consolidations, a sign of acute respiratory distress syndrome (ARDS) which may put the patient on a ventilator and could be the cause of death [7]. However, aggravated symptoms with respiratory complications have been frequently observed in many younger and healthy patients as well. As the pathology is quite recent, there is awfully little insight into other risk factors and into the cause of disease progression that is observed only in some cases and not in others.

Similar to tuberculosis (TB), chest radiography, in the case of Covid-19 may not just be a potential diagnostic tool, but also can support in assessing the chosen treatment, evaluating the trajectory of disease progression as well as in predicting outcomes. X-rays are cost-effective, quick and widely available even in rural areas and hamlets. Moreover, the radiography reports can be automated using computational Artificial Intelligence (AI) based algorithms that can facilitate immediate diagnosis, further reducing the burden on the healthcare systems and facilitating diagnosis in remote areas with no dedicated radiologist. A plethora of emerging work from the past two months has illustrated that deep learning (a state of art AI technology) has the capability to delineate Covid-19 pathology from pneumonia [8] as well as TB [9] and from healthy subjects [10]. Majority of the works have proposed AI based techniques for diagnosis of Covid-19 from radiographs [11] or from chest CT scans [5-11] and quote that such screening may detect Covid-19 with a 97% sensitivity whereas viral PCR, the state of art technique is upto 72% sensitive [12]. Such high sensitivity, although is attractive, many studies illustrate this have a patient selection bias, where the chest radiographs have been taken after the disease progressed. Nonetheless, the importance of AI based prescreening is turning out to be a crucial tool as it takes a few seconds whereas the final diagnostic report may take a few minutes to hours and is dependent on various factors, especially related to the increasing patient volumes. AI prescreening can substantially help in busy radiology departments to optimize workflows. This aspect must be given utmost importance in the prevailing pandemic circumstances as instant identification can be significantly effective for triaging patients, this can support subsequent treatment planning as well as disease containment. Chest radiography with AI can therefore be a remarkable auxiliary diagnostic tool that can prescreen and isolate Covid 19 suspected patients promptly whilst RT-PCR substantiates.

This work proposes an automated deep learning-based framework with rapid report generation for delineating Covid-19 and TB in two different settings from their initial chest x-ray. Though most of the solutions focus on AI pre-screening and triage, we focus on using an AI embedded end to end imaging platform, which assists imaging experts from using the benefits of prescreening and provide structured radiology report and also use the powerful analytic tool to understand day to day workflow better. AI infused platform with AI embedded in the medical image analysis and workflow can make solutions robust, streamline radiology workflows and assist in reducing imaging experts’ burnouts. They can be successfully integrated with existing hospital Picture Archival and Communication System (PACS), Radiology Information System (RIS) and Hospital Information System (HIS). Hospitals which do not have these solutions can use the AI platform as the primary PACS. The platform has been earlier extensively tested for tuberculosis and pneumonia and other chest ailments with a unique model of “AI with an expert in the loop”.

The model has dual advantages the AI assists the imaging experts in performing a quick pre-screening and provide an auto-generated report. The medical imaging expert uses the report as the base report and can add/ edit and submit this report which can significantly reduce the efforts. The report given by expert is taken as a feedback to ensure that AI models gets consistently trained and retrained and more robust with inputs from the experts. With new forms of community acquired pneumonia like Covid-19, the importance of such a framework had now been adapted to incorporate quick diagnosis of Covid-19 from chest radiographs. Here we present a comprehensive validation of the AI based chest -x-ray screening on 9098 radiographs in a hospital-based setting, out of which around 504 patients with RT-PCR validations. We also present another validation of use of this tool for a large-scale TB population screening program involving over 80,000 subjects with the combination of AI with experts in the loop. We believe that the validations can pave the way for using the AI infused platform for handling large datasets in busy hospital and ICU settings on one side and also support village to village chest x-ray screening in mobile van screening for TB diagnosis. This will not only help diagnosing the current pandemic of Covid-19 but also will identify patients with TB consequently supporting the eradication of both the diseases expeditiously.

## Methods

### Dataset 1

This was a retrospective study that was performed in Nanavati Hospital, Mumbai. The analysis included 9098 chest radiographs belonging to 3180 patients. The RT-PCR test results on various dates were available for 604 patients accounting for a total of 1656 radiographs. Out of these 604 patients, 484 tested positive while the rest 120 tested negative.

### Dataset 2

We collaborated with the Clinton Health Access Initiative (CHAI) and a large City Corporation under the TB Free Cities initiative to use AI in a population screening project in India. The Corporation sends vans equipped with X-ray machines to regions with priority population groups. We reported digital chest X-ray scans of screened patients. These scans were evaluated by qualified radiologists who labelled each scan as TB-positive or TB-negative. For all TB-positive images, bounding boxes were drawn around the region in the image suspected to contain the infection.

The presence of one or more of the following conditions was considered an indication of TB for marking a scan as positive: nodular shadows, infiltrates, pleural effusion, pneumonia or consolidation-like features, fibrosis, pleural thickening, granuloma, bronchiectasis, scarring, lymph node, and calcified pleural plaques. From June 2019 to December 2019, we received about 7000 chest X-ray scans at a specific time interval in batches. During this period, the models were retrained on every successive batch using radiologist annotations. This led to a continuous improvement in model performance. The final models were trained on data up to December 2019 consisting of 42,837 images.

### Platform

Both the datasets were analyzed using the Genki Platform from DeepTek Inc that was deployed in Nanavati hospital (dataset 1) and in a mobile diagnostic van (dataset 2). The Genki platform is a secure and compliant web-based framework that includes an AI based tool for prescreening and diagnosing the underlying chest pathology and rapidly generating the radiology report. It can either be used in a completely automated fashion (in places where radiologist is unavailable as a prescreening tool) or with a radiologist in loop (expert in the loop) where the expert can confirm or edit the AI markings and analysis. In both the modes, diagnostic time is drastically reduced as majority of the situation the AI facilitates instant diagnosis and in addition the reporting is quick as the radiologist does not need to dictate or type the report. The AI inputs leads in generated structured radiology reports making the job of the imaging experts faster. Moreover, the tool also generates class activation maps to delineate the pathology, standardizing the radiology workflow allowing to create structured and quantified radiology reports. The internal structure of the Genki platform includes two separate deep learning-based modules: one for classification and other for segmentation (see Figure 2). Figure 1 demonstrates the platform display where the AI automatically detects the abnormality on the scan and provides the probable diagnosis in the lower taskbar (circled in orange). For example, in the first row, the diagnosis given by AI is tuberculosis while in the second row the diagnosis is Covid-19. The report generation is done automatically as shown in the image.

**Figure 1:**
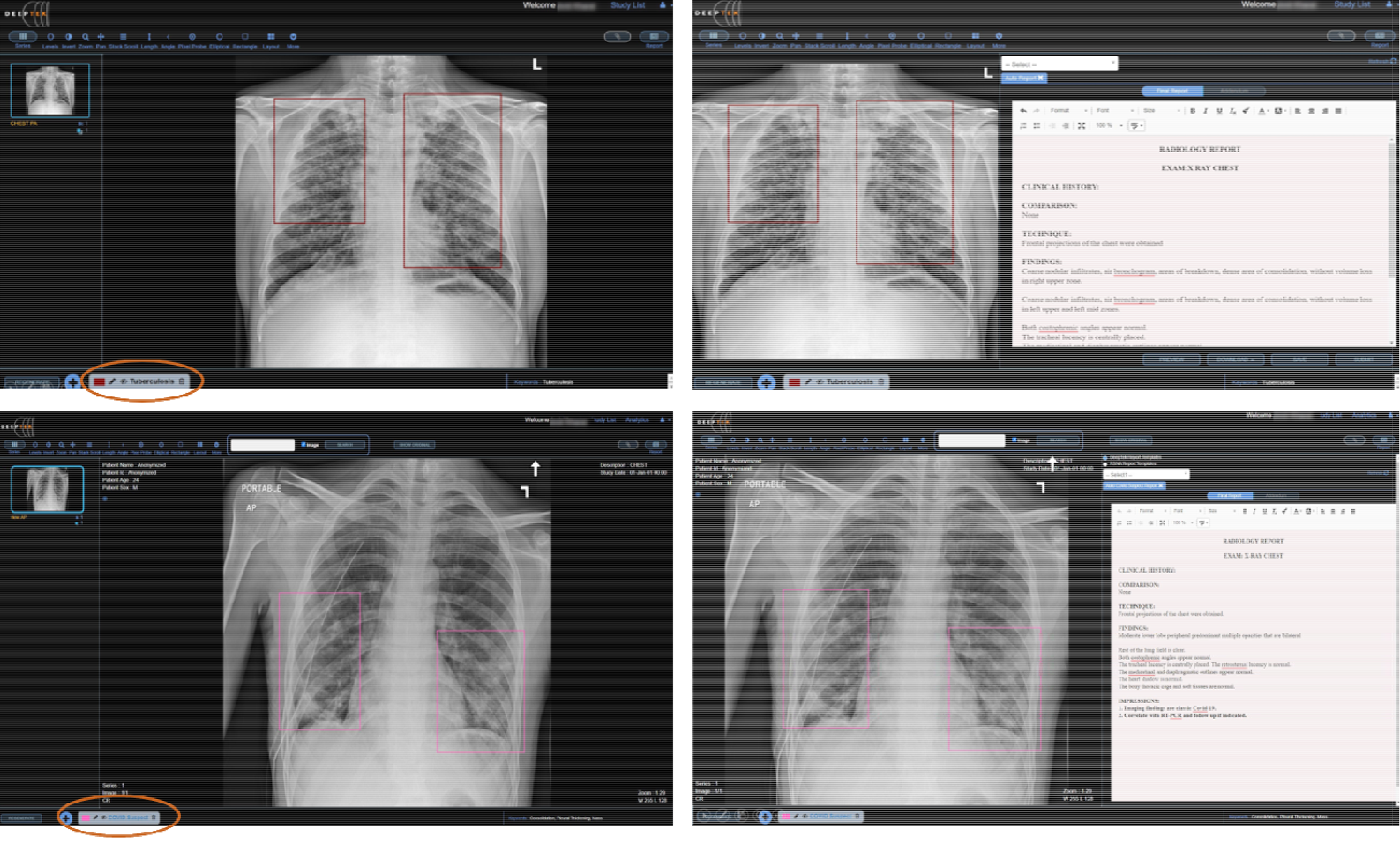
Genki platform by Deeptek Inc, where the chest x-ray is annotated and diagnosed using AI. Moreover, the structured semi-automated report generation is showcased for TB and suspected Covid infection. The platform creates an efficient radiology workflow with quick prescreening and rapid report generation by experts.

**Figure 2:**
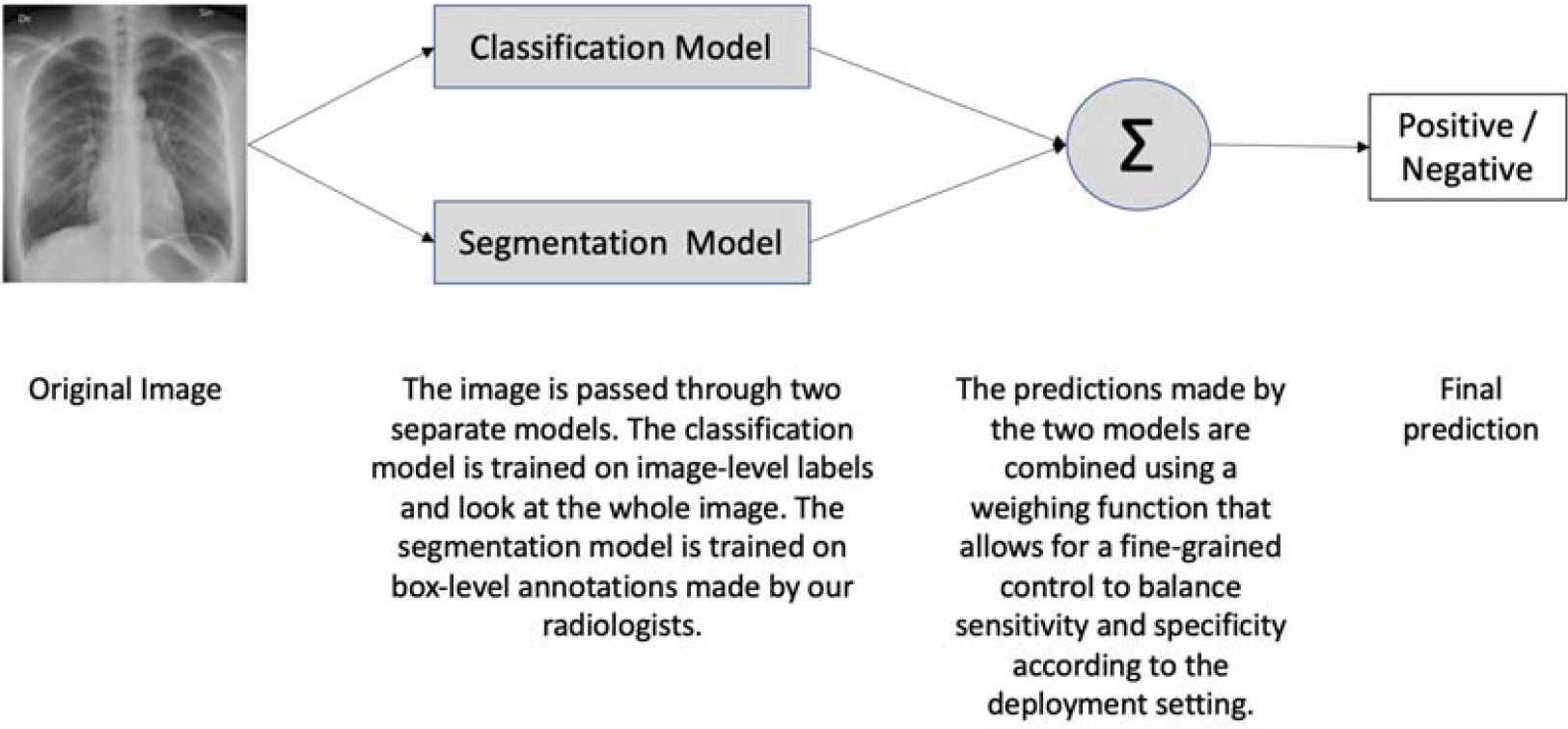
Architecture of DeepTek model pipeline for Covid-19 and TB prescreening. The outputs from the classification and segmentation models are combined using a weighing function to enable fine-grained control over balancing sensitivity and specificity according to the setting.

**Figure 3:**
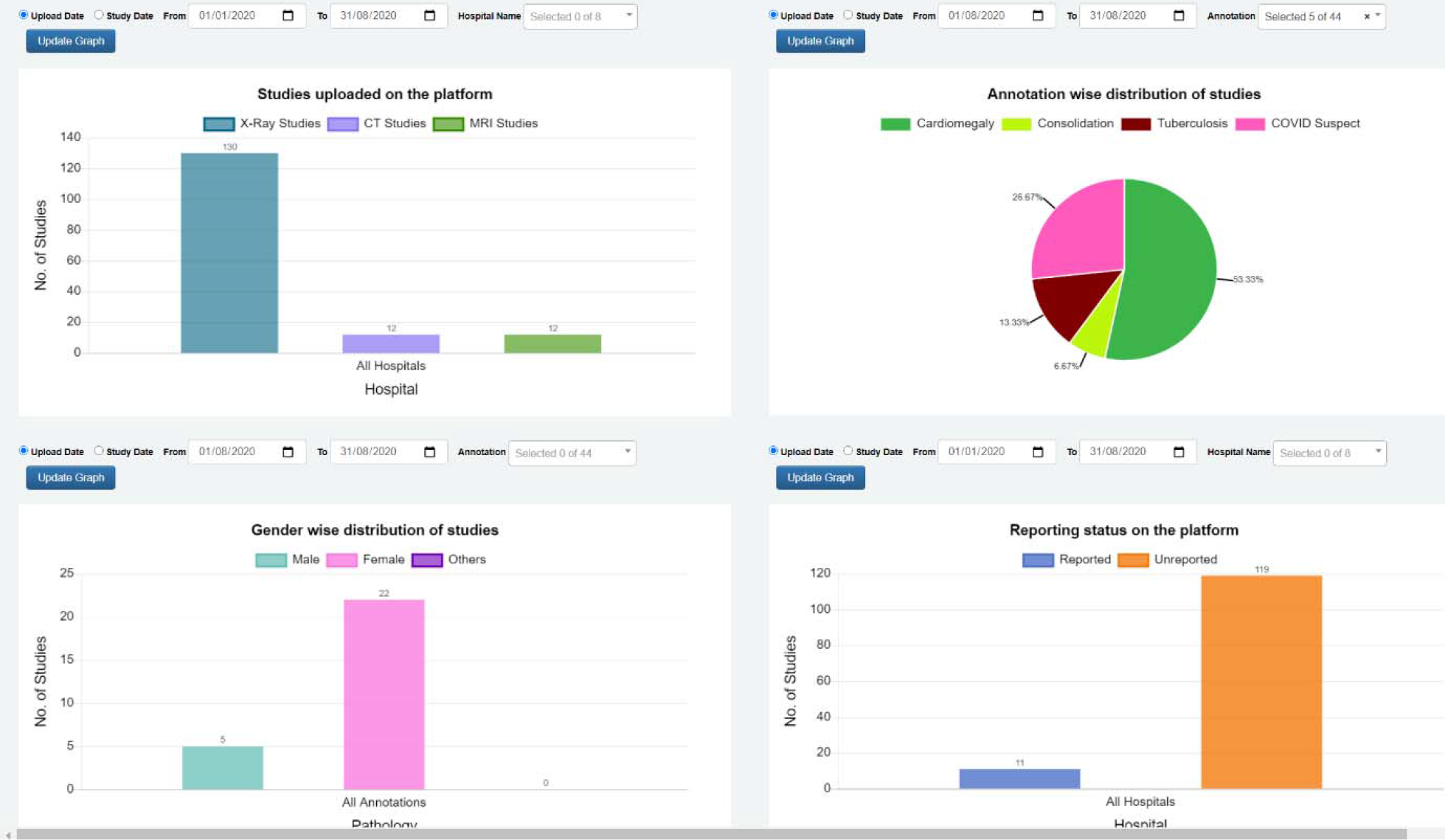
Analytics Dashboard on the Genki platform (DeepTek Inc) for TB and Covid Screening. In a clockwise manner showcases – a] annotation usage metrics for x-ray studies, b] report pending status on the platform, c] gender wise distribution of pathology and d] total number of x-rays, CT and MRI on platform.

### Test Analysis

For Covid-19 dataset, we first validated the AI model on one chest radiograph per patient, this was acquired between day 1-5 after the patients RT-PCR was positive (design 1)). This validation was crucial to check if the Genki platform was able to capture and identify the underlying Covid-19 pathology and delineate it from normal scan and other chest abnormalities. However, this design does not guarantee the diagnostic ancillary utility of the tool as the chest scan could illustrate abnormalities on day 5 that were not visible on the day the RT-PCR was performed. To alleviate this issue, in design 2, we chose only the radiographs that were acquired the day before or on the day the RT-PCR was performed. The radiographs were analyzed using the platform as well as annotated by a radiologist for validation. For multiple radiographs of the same patient we combined the annotations and predictions using an OR operation. The design 1 and 2 details are presented in Table 1. In both designs, the results were quantified as a match or non-match between diagnosis of radiologist and RT-PCR results.

**Table 1:** Detailed Explanation of the experimental design for Covid-19 data.

|  | Design-1 | Design-2 |
| --- | --- | --- |
| **Sample** | One patient was considered as one sample | One (X-ray, PCR) pair where X-ray was taken one day before, on same day, or one day after the PCR test was considered as one sample |
| **Sample considered ground-truth positive** | If any one of the many RT-PCR tests was positive, sample was considered positive. | If PCR test was positive, sample was considered as positive |
| **Samples considered ground-truth negative** | If none of the many RT-PCR tests were positive, sample was considered negative. | If PCR test was negative, sample was considered as negative |
| **Sample predicted as positive** | If any one of the many X-ray images were predicted as positive, sample was considered positive. | If PCR test was positive, sample was considered as positive |
| **Sample predicted as negative** | If none of the many X-ray images were predicted as positive, sample was considered negative. | If PCR test was negative, sample was considered as negative |

For the TB dataset, the experiment design is as per table 2.

**Table 2:** Detailed Explanation of the experimental design for TB screening.

| Experiment Design for TB |  |
| --- | --- |
| Studies | All studies were digital chest X-rays taken from portable units installed in mobile vans |
| Ground Truth | Ground truth for developing models was annotations made by qualified radiologists |
| Pathologies Considered for Marking X-Ray as TB-positive | Infiltrates, Nodular shadows, Cavitation, Breakdown, Lymph Nodes, Pleural Effusion, Bronchiectasis, Fibrosis, Scar, Granuloma, Nodule, Pleural Thickening, Calcification, Calcified Lymph Nodes, Calcified Pleural Plaques |
| Annotations | Boxes were drawn around the infected region |
| Models | Classification and segmentation models were built |
| Evaluation | Final model was evaluated prospectively to determine how well it could diagnose TB from the input X-ray |

The AI triage results were reviewed by an imaging expert in the loop and further validated to prepare a final diagnostic report within 60 minutes of the scan. The AI triage happened within few seconds on receipt of the digital x-ray on the platform from the x-ray modality with which it was configured. The model assessed all the radiographs of the patients and classified each of them into one of two classes: (1) patient likely to have TB and (2) patient unlikely to have TB. All these images were retrospectively evaluated by an expert radiologist.

The radiographic evaluation with AI also allowed us to quantify in symptomatic and those with comorbid conditions the amount of lung involvement. Unlike RT-PCR where results are seen as positive and negative, the lung quantification on X-ray imaging and correlation with comorbidity allows for better decision making in terms of allocating valuable resources like admission in hospital in low resource settings.

## Results

As illustrated in table 3 for Covid 19 screening, the AI based platform approaches human imaging experts level output when compared to radiologists’ annotations as ground truth. The model demonstrated a sensitivity of 0.67, 95% CI [0.59, 0.74], at a specificity of 0.43, 95% CI [0.26, 0.41], with an accuracy of 0.46, 95% CI [0.41, 0.52] when compared to RT-PCR. However as described above when compared to imaging experts (radiologists) the sensitivity and specificity was far better. The AI models matched radiologist’s performance in diagnosis of Covid 19 from chest x-rays and obtained a sensitivity of 0.87 at a specificity of 0.60 when compared to radiologists as ground truth.

**Table 3:** Performance results of Radiologist and AI models vs RT-PCR results as ground truth for Covid 19 screening.

|  | Design-1 |  |  |  | Design-2 |  |  |  |
| --- | --- | --- | --- | --- | --- | --- | --- | --- |
|  | Radiologist | 95% CI | AI | 95% CI | Radiologist | 95% CI | AI | 95% CI |
| **Sensitivity** | 0.32 | [0.28, 0.36] | 0.66 | [0.62, 0.7] | 0.38 | [0.31, 0.46] | 0.67 | [0.59, 0.74] |
| **Specificity** | 0.36 | [0.27, 0.45] | 0.32 | [0.24, 0.40] | 0.52 | [0.47, 0.57] | 0.43 | [0.26, 0.41] |
| **Precision** | 0.67 | [0.61, 0.73] | 0.8 | [0.76, 0.83] | 0.27 | [0.22, 0.33] | 0.36 | [0.3, 0.41] |
| **Accuracy** | 0.33 | [0.29, 0.37] | 0.59 | [0.55, 0.63] | 0.47 | [0.43, 0.52] | 0.5 | [0.46, 0.55] |
| **F1** | 0.44 | [0.39, 0.48] | 0.72 | [0.69, 0.75] | 0.32 | [0.26, 0.38] | 0.46 | [0.41, 0.52] |

| Batch | Modality | Evaluator | Sensitivity | Specificity | Sample Size |
| --- | --- | --- | --- | --- | --- |
| 1 | X-ray | Radiologist-1 | 0.4 | 0.52 | 460 |
| 1 | X-ray | Radiologist-2 | 0.42 | 0.5 | 460 |
| 1 | X-ray | AI | 0.63 | 0.44 | 460 |
| 1,2 | X-ray | Radiologist-1 | 0.38 | 0.52 | 502 |
| 1,2 | X-ray | AI | 0.67 | 0.43 | 502 |

As illustrated in table 5, the AI model for TB demonstrated a sensitivity of 0.89, 95% CI [0.87, 0.91], specificity of 0.86, 95% CI [0.85, 0.86], and accuracy of 0.86, 95% CI [0.85, 0.86] when evaluated retrospectively on a hold-out test set. When evaluated prospectively on live on-field data, it demonstrated a sensitivity of 0.91, 95% CI [0.88, 0.93], specificity of 0.82, 95% CI [0.81, 0.82], and accuracy of 0.82, 95% CI [0.81, 0.83].

**Table 4:**
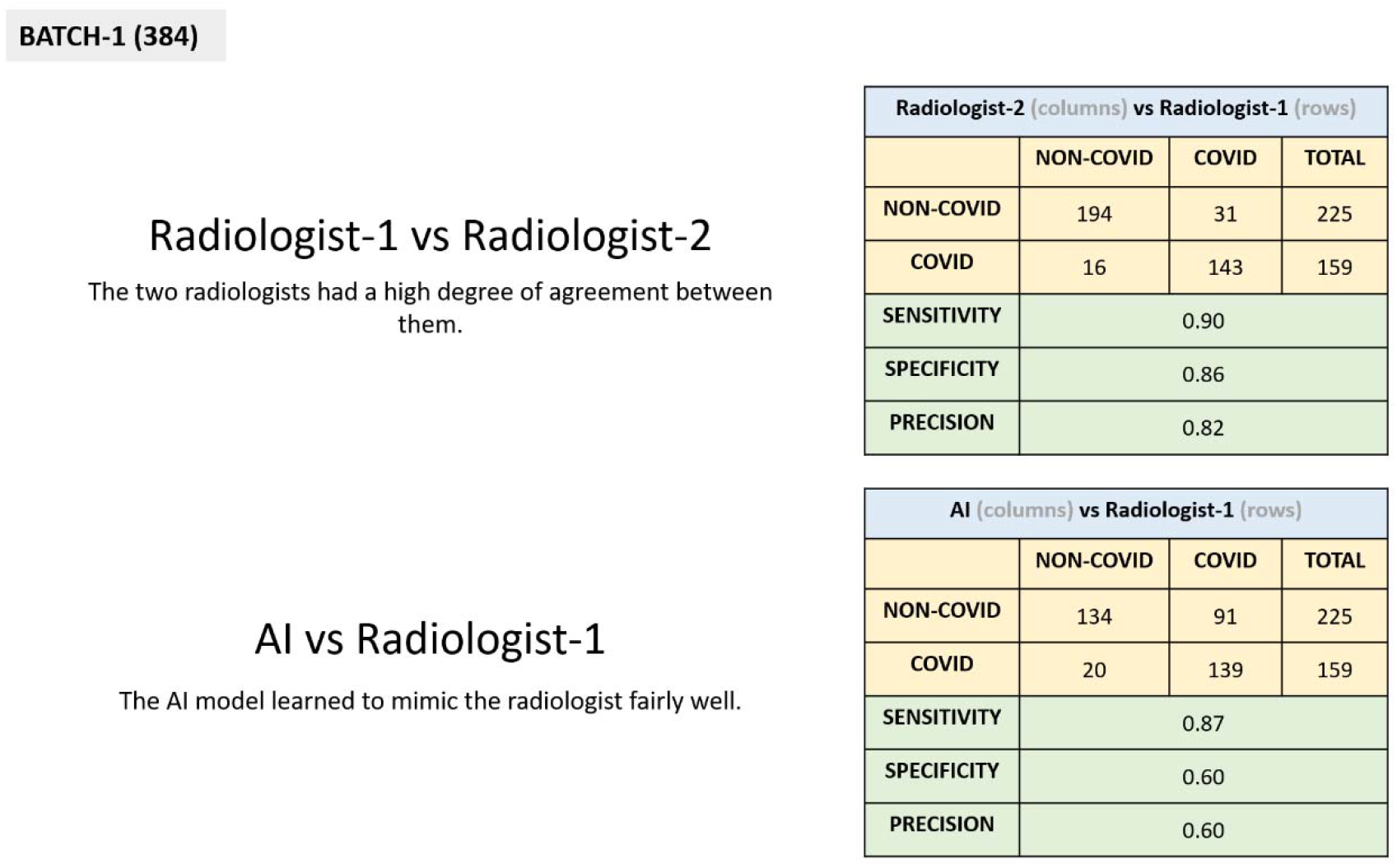
BATCH-1 (384) Radiologist-1 vs Radiologist-2 The two radiologists had a high degree of agreement between them.

| Radiologist-2 (columns) vs Radiologist-1 (rows) |  |  |  |
| --- | --- | --- | --- |
|  | NON-COVID | COVID | TOTAL |
| NON-COVID | 194 | 31 | 225 |
| COVID | 16 | 143 | 159 |
| SENSITIVITY | 0.90 |  |  |
| SPECIFICITY | 0.86 |  |  |
| PRECISION | 0.82 |  |  |

**AI vs Radiologist-1** The AI model learned to mimic the radiologist fairly well.
| AI (columns) vs Radiologist-1 (rows) |  |  |  |
| --- | --- | --- | --- |
|  | NON-COVID | COVID | TOTAL |
| NON-COVID | 134 | 91 | 225 |
| COVID | 20 | 139 | 159 |
| SENSITIVITY | 0.87 |  |  |
| SPECIFICITY | 0.60 |  |  |
| PRECISION | 0.60 |  |  |

**Table 5.** Performance results of Radiologist and AI model’s vs Radiologist annotations results as ground truth for TB screening.

| Performance evaluation of TB models |  |  |
| --- | --- | --- |
|  | Retrospective Evaluation on<br>June 2019 – Dec 2019 data | Prospective Evaluation on<br>Jan 2020 to Jun 2020 Data |
| Sensitivity | 0.89, 95% CI [0.87, 0.91] | 0.91, 95% CI [0.88, 0.93] |
| Specificity | 0.86, 95% CI [0.85, 0.86] | 0.82, 95% CI [0.81, 0.82] |
| Accuracy | 0.86, 95% CI [0.85, 0.86] | 0.82, 95% CI [0.81, 0.83] |

Our models achieved expert level performance in diagnosing both COVID-19 and TB from chest radiographs.

## Discussion

The work demonstrated that AI infused platforms have a wide applicability in diagnosing chest anomalies including TB and Covid-19 with sensitivity and specificity matching imaging experts’ evaluations. Moreover, for the Covid-19 data we also illustrated that regardless of the study designs the sensitivity was higher than manual identification.

Despite the low sensitivity of RT-PCR of around 35 to 71% it is still considered to be the gold standard diagnosis for Covid-19 [13]. Nevertheless, RT-PCR tests are costly and laborious and require an elaborate lab setup with expert technicians which could be limiting and prohibitive in a pandemic situation. It is therefore crucial to investigate and have parallel auxiliary track for diagnosis in symptomatic so as to isolate probable suspects and for timely treatment that will consequently reduce the burden on healthcare bringing the situation under control.

Alternative strategies may include CBC in symptomatic cases based on the presence of neutrophilia and leukopenia. However, this is largely not enough as it lacks specificity. SPO2 monitoring is also crucial. Falling SPO2 levels is also considered to be critical and indicates higher comorbidity status. The other investigation in line for symptomatic could be a simple digital chest x-ray. Primarily, the use of chest x-ray can decide whether or not the patient needs home or hospital quarantine. X-rays are also broadly available, highly portable, even available as hand held, even battery operated, relatively cheap and often done free of cost under various schemes in most countries. India has a well-established network of X-ray centers that are largely spread across the nation, this network has been catering to TB screening program and can be repurposed for Covid-19 screening. Most other countries too similarly have a huge availability of X-ray machines and herein lies the strength of incorporating x-ray into screening for Covid-19.

On a broad scale x-ray screening for symptomatic alone may not be exciting. The reason could be multiple - the patches of Covid-19 may be subtle and difficult to assess in patients who are not symptomatic. The patches are not consistent in terms of site and size and X-Ray as a modality may have inherent flaws to diagnose these subtle nuances and hence is not very specific. There may be many false positives [14]. The classic pathognomic features of Covid-19 pneumonia pattern may be difficult to assess if there is motion blur or quality issues like a mid-expiratory film. However corrective measures exist, such as adjusting the threshold of the algorithm can be done to reduce false positives. Nonetheless, x-rays are a part of the investigation for screening individuals to undertake decisive actions such as factoring whether suspected patients need self-isolation at home or hospital quarantine. These decisions are important because every patient admitted in the current scenario will stretch the healthcare resources thin.

Advanced economies due to the high density of CT scanners, are capable of scanning an immense volume of patients with CT, and this has largely enabled them to assess and quantify pneumonia particularly that due to Covid-19; Covid-19 pneumonia pattern is pathognomonic. However, even the best of imaging available to us cannot establish a final diagnosis as other community acquired pneumonia can mimic Covid 19 pattern. (Gene expert test still remains the gold standard).

Understanding the TB diagnostics paradigm - In the last few months the biggest casualty due to the Covid 19 pandemic has been TB screening programs. These programs have almost come to a standstill and have a massive impact on people affected by TB, especially among the most vulnerable who are struggling to get their treatment, care and other types of support [15].

The way forward could be to improve active screening through clinical examination and screening chest x-rays for symptomatic and active surveillance. There have not been any significant pilots due to Covid 19 but this should be the key moving ahead. If there is a value add happening through population screening for TB in addition to Covid 19 this could be a key to tackle two chest conditions in one go. The tools can also be optimized for lung cancer and lung nodule screening, quantification and follow up assessment. Considering the uncertainty of Covid 19 pandemic timelines it is crucial to move ahead with other screening programs with protocols.

Considering both are primarily (Covid 19 and TB) involving chest, both are respiratory illnesses and share an overlapping symptomatology, presenting with fever, cough, but then that’s where the similarity ends [16][17]. Both have vastly different timelines - acute presentation in Covid 19 and Tuberculosis where it typically spans weeks.

The problems caused by COVID-19 infection in a population already burdened with TB are multi-faceted. TB programs globally have hit a road block due to the emergence of COVID-19. The first major challenge these patients face is a social stigma as cough has become synonymous with COVID-19 for the general public. The second challenge is the lack of accessibility to diagnostics, as the supply chains have been interrupted due to lockdowns globally resulting in manufacturing and logistical issues.

The way forward could be to improve active screening through clinical examination and screening chest x-rays for the symptomatic. There have not been any significant pilots to underscore the presence of COVID-19 in surveillance settings but this should be the key moving ahead. If there is a value add happening through population screening for TB plus symptomatic Covid-19 this could be a key to tackle two chest conditions in one go. However, it is an exercise which needs to be done.

Current technology and AI models can screen for multiple pathologies, apart from TB and COVID-19 such as pleural effusion, consolidation, atelectasis, lung mass, lung nodule, atelectasis, lines, tube, pneumothorax, opaque hemithorax, cardiomegaly, hernias. The AI has broadly helped to screen and triage all the above conditions with ease apart from emphysema and acute conditions like pneumothorax. Using screening solutions can be cost effective especially in hot spots and can be considered as a complementary test to the ongoing tests. Whatever ways and means available to improve diagnosis, triage of studies should be used to limit the disease and its spread and meet targets for disease eradication.

Chest x-ray screening in individuals with comorbidities is also crucial to identify those at risk especially at hot spots. Screening them at or close to home is also the key to limit the movement of a large number of patients to the hospital [18]. Portable x-ray mounted in a van seems the best solution and it should be explored further in greater depth as it opens up avenues for instantaneous triage utilizing AI for pre-screening, further backed by radiology experts in the loop. The key advantage is instant triage of these patients within seconds of the x-ray. The studies which show the evidence of a patch or similar pathology should be isolated at least until an expert radiologist in the loop reviews the scan and gives a final diagnosis. On the flip side there are limitations in the X-ray examination, COVID 19 carriers and asymptomatic may not be diagnosed. Also, there is risk of exposure to radiation although minimal. These can be subjected to surveillance testing by RT-PCR and serology through a decision support tool as the one provided by British Society of Thoracic Imaging [19]. This may also help bring down the burden of the tests to be performed using RT PCR [20].

Although, using RT-PCR has led to a few false-negative results they have been attributed to the quality of the kit, the collected sample, or performance of the test. RT-PCR has been widely deployed in diagnostic virology and has yielded few false-positive outcomes as well [21]. In conclusion, However the dynamics of diagnostics are changing on a day to day basis and are dependent on various factors and the foremost being access to healthcare.

Performing clinical assessment, SPO2 monitoring, CBC and digital Xray of the chest in symptomatic to assist triage could be an option which can help tide the crisis of Covid-19 and can be decided on the available infrastructure and guidelines can be structured accordingly. Promoting mobile van diagnostic testing can help tackle especially in hot spot two conditions – Covid 19 and TB [22]. This could also be an opportunity in disguise to scale the TB screening process, if India want to meet the target of TB eradication by 2025 and WHO wants to adhere to the global target of TB elimination by 2030.

Our experience in screening of TB also shared some other tangible benefits such as AI also assisted in triage of other pathologies such as community acquired pneumonias. nodules, scar, pleural effusion, pleural thickening and cardiomegaly. The AI has broadly helped to screen and triage all the above conditions with ease apart from emphysema and acute conditions like pneumothorax. Using screening solutions can be cost effective especially in hot spots and can be considered as a complementary test to the ongoing tests.

## Summary

To summarize we present two conditions: a rapidly spreading global pandemic of COVID 19 and other end TB a scourge on mankind for 15,000 years. Both have predominant lung involvement, though not limited to it.

AI based instant triage and prescreening offers hope to optimize workflows in radiology department, enable quick prescreening, generate structured radiology reports, allow focused utilization of scare and costly resources and use the powerful dashboard for disease insights. The platform can serve dual purpose like prescreening for COVID-19 and TB and provides instant triage especially in situation of low resources. Both the systems can be deployed together or can be deployed standalone.

By creating automated structured reporting, the system can help reduce effort further and create a methodical reporting system which can be customized by the organization to their requirements.

Both the studies showed that the AI output matched the output of expert radiologists and this indicates that AI can assist an expert radiologist in prescreening and support structured reporting initiatives and instant triage functionalities. Both these features are important for prompt and efficient treatment so that the patients are not lost for follow up. The model can be further fine-tuned by adjusting thresholds to support these activities and by either reducing the number of false positives or negatives. The faster organizations and public screening programs adopt AI for triage, structured reporting using smart reporting tools and a combination of AI based image analysis, AI embedded workflows, the faster it will be to possible to scale these “AI plus experts in the loop” design system to make healthcare accessible and available.

## Data Availability

This has been approved by Institutional Review Board at Nanavati Hospital and its ethics committee and we have now received the letter which has been provided by the ethics committee. We can share the attachment of the same.

